# Multimorbidity Patterns and Socioeconomic Determinants in a resource-limited setting: A Clustering Analysis

**DOI:** 10.1101/2025.10.15.25337896

**Authors:** Olalekan A. Uthman, Matthew Hazell, Muhammed Mubashir Babatunde Uthman, Kolawole W Wahab, Ponnusamy Saravanan, Andre Pascal Kengne, Paramjit Gill

## Abstract

**Importance:** Multimorbidity, the coexistence of multiple long-term conditions, is a growing public health challenge in South Africa. Understanding the patterns of multimorbidity and their socioeconomic determinants is crucial for developing prevention and control solutions.

**Objective:** To identify and characterize multimorbidity clusters and their socioeconomic determinants in the South African population using data from the Demographic and Health Survey (DHS).

**Design, Setting, and Participants:** This cross-sectional study used data from the recent South Africa DHS, a nationally representative household survey. The study included 5,342 individuals aged 18 years and above who participated in the adult health module of the survey. Data were collected through interviews and biomarker measurements between June and November 2016.

**Main Outcome and Measures:** The primary outcome was multimorbidity, defined as the presence of two or more chronic conditions. Twelve chronic conditions were considered: tuberculosis, hypertension, stroke, high blood cholesterol, anaemia, chronic bronchitis, diabetes, asthma, cancer, heart disease, HIV, and chronic pain. Socioeconomic determinants included wealth index, education level, occupation, health insurance, marital status, age, sex, ethnicity, and media access.

**Results:** Of the 5,342 participants, 2,382 (44.6%) had multimorbidity. Four distinct multimorbidity clusters were identified: “Low Morbidity Group” (low prevalence of chronic conditions), “Cardiometabolic Cluster” (high prevalence of hypertension and diabetes), “Chronic Infectious Disease Cluster” (high prevalence of tuberculosis and HIV), and “Complex Chronic Disease Cluster” (high prevalence of multiple chronic conditions, including cancer, stroke, and heart attack). Multinomial logistic regression analysis revealed socioeconomic disparities in multimorbidity patterns, with lower levels of education, unemployment, and poverty associated with membership in the clusters characterized by a higher burden of chronic diseases.

**Conclusions and Relevance:** This study identified four distinct multimorbidity clusters in the South African population, each characterized by unique patterns of chronic disease co-occurrence and socioeconomic determinants. The findings highlight the need for tailored interventions and policies that address the specific needs of each multimorbidity cluster while also tackling the underlying social and economic determinants of health.

## Introduction

Multimorbidity, the coexistence of two or more chronic conditions in an individual, has emerged as a significant public health challenge in low- and middle-income countries (LMICs), including South Africa(1). The rising prevalence of multimorbidity in these settings is driven by a complex interplay of factors, such as rapid urbanization, lifestyle changes, population aging, and the double burden of communicable and non-communicable diseases(2, 3). The impact of multimorbidity on individuals, healthcare systems, and societies is substantial, leading to poorer health outcomes, reduced quality of life, increased healthcare utilization and costs, and exacerbated health inequities(4, 5). Despite the growing recognition of multimorbidity as a global health priority, there is a paucity of research on the patterns and determinants of multimorbidity in LMICs, particularly in sub-Saharan Africa(6). Existing studies have primarily focused on the prevalence of individual chronic conditions or the overall burden of multimorbidity, with limited attention to the heterogeneous patterns of disease clustering and their socioeconomic determinants(7). Understanding these patterns and determinants is crucial for designing targeted interventions and policies to prevent and manage multimorbidity effectively.

Recent advances in data-driven approaches, such as unsupervised machine learning, offer novel opportunities to uncover the complex patterns of multimorbidity and identify distinct subgroups of individuals with similar disease profiles(8). These approaches can help to move beyond the traditional single-disease paradigm and provide a more nuanced understanding of the multimorbidity landscape. Furthermore, investigating the socioeconomic factors associated with multimorbidity clusters can shed light on the underlying health inequities and inform the development of tailored interventions for vulnerable populations(9).

This study aims to address the knowledge gaps in understanding the patterns and socioeconomic determinants of multimorbidity in South Africa, using data from the Demographic and Health Surveys (DHS). By employing unsupervised machine learning techniques, such as clustering algorithms and network analysis, this study will provide a comprehensive characterization of multimorbidity clusters and their associated socioeconomic and environmental factors. The findings of this study will have important implications for public health policy and practice in South Africa and other LMICs facing similar challenges. Identifying the distinct multimorbidity clusters and their socioeconomic and environmental determinants will inform the development of targeted prevention and management strategies, resource allocation, and health system strengthening efforts to address the growing burden of multimorbidity(10). This study will contribute to the growing body of evidence on the social determinants of health and the need for intersectoral collaboration to tackle health inequities(11).

## Methods

### Study design and data source

This study employed a cross-sectional design using data from the 2016 South Africa Demographic and Health Survey (SADHS). The SADHS is a nationally representative household survey that provides comprehensive information on various health indicators, including chronic conditions and socioeconomic factors. The survey followed a stratified two-stage sampling design, with households selected from primary sampling units (PSUs) based on the 2011 census enumeration areas.

In the first stage, 750 PSUs were selected with probability proportional to size, stratified by urban, rural, and traditional areas within each of the nine provinces in South Africa. In the second stage, a fixed number of 20 dwelling units (DUs) were randomly selected from each PSU, resulting in a total sample size of 15,000 DUs. All household members in the selected DUs were eligible for the primary modules on women, fertility, and children, while half of the DUs were subsampled for the adult health module, which included information on chronic conditions and biomarker collection.

Trained interviewers administered the questionnaires using computer-assisted personal interviewing systems. Biomarker measurements, including anthropometry, blood pressure, and blood samples for HIV and anaemia testing, were collected by trained health personnel.

### Study population and variables

The study population included men and women aged 18 years and above who participated in the adult health module of the SADHS. Individuals with missing data on the multimorbidity outcome were excluded from the analysis. The primary outcome variable was multimorbidity, defined as the presence of two or more chronic conditions in an individual. Twelve chronic conditions were considered in this study:

1. Tuberculosis: Participants were asked if they had been diagnosed with tuberculosis by a healthcare professional within the past 12 months.
2. Hypertension: Blood pressure measurements were taken using a digital blood pressure monitor, and participants were classified as hypertensive if they had a systolic blood pressure ≥140 mmHg or a diastolic blood pressure ≥90 mmHg, or if they reported current use of antihypertensive medication.
3. Stroke: Participants were asked if they had ever been diagnosed with a stroke by a healthcare professional.
4. High blood cholesterol: Participants were asked if they had ever been diagnosed with high blood cholesterol by a healthcare professional.
5. Anaemia: Haemoglobin levels were measured using a HemoCue analyser, and participants were classified as anaemic if their haemoglobin levels were <12 g/dL for women and <13 g/dL for men.
6. Chronic bronchitis: Participants were asked if they had been diagnosed with chronic bronchitis by a healthcare professional.
7. Diabetes: Blood glucose levels were measured using a point-of-care device, and participants were classified as diabetic if their fasting blood glucose was ≥126 mg/dL or if they reported current use of insulin or oral hypoglycaemic medication.
8. Asthma: Participants were asked if they had ever been diagnosed with asthma by a healthcare professional.
9. Cancer: Participants were asked if they had ever been diagnosed with any type of cancer by a healthcare professional.
10. Heart disease: Participants were asked if they had ever been diagnosed with heart disease by a healthcare professional.
11. HIV: Blood samples were tested for HIV using the Determine HIV-1/2 rapid test, and positive results were confirmed using the Uni-Gold rapid test.
12. Chronic pain: Participants were asked if they had experienced persistent pain lasting more than three months.

Explanatory variables included socioeconomic factors, individual-level health variables, access to media, and demographic factors. Socioeconomic factors included wealth index (poorest, poorer, middle, richer, richest), education level (no education, primary, secondary, higher), occupation (not working, manual, non-manual), health insurance (yes, no), and marital status (never married, currently married, formerly married). Individual-level health variables included body mass index (underweight, normal, overweight, obese), dietary health (poor, moderate, good), sugary drink intake (none, 1-6 times per week, daily), smoking status (never, former, current), alcohol consumption (never, occasional, regular), and exposure to smoke at work (yes, no). Access to media was assessed using two variables representing access to old media (radio, television) and new media (internet, social media). Demographic factors included age (18-24, 25-34, 35-44, 45-54, 55-64, ≥65 years), sex (male, female), and ethnicity (Black African, White, Coloured, Indian/Asian).

### Data analysis

#### Stage 1: Cluster identification

Unsupervised machine learning techniques, specifically K-means clustering, were employed to identify distinct multimorbidity clusters. K-means clustering aims to partition the dataset into K clusters, where each data point belongs to the cluster with the nearest mean. The algorithm iteratively assigns data points to clusters and updates the cluster means until convergence.

The input features for the clustering algorithm included the binary variables indicating the presence or absence of each of the 12 chronic conditions, along with the explanatory variables (socioeconomic factors, individual-level health variables, access to media, and demographic factors). Before clustering, the data were pre-processed by handling missing values (using multiple imputation), scaling the continuous variables, and one-hot encoding the categorical variables. The optimal number of clusters (K) was determined using a combination of the elbow method and silhouette analysis. The elbow method involves plotting the within-cluster sum of squares (WCSS) against the number of clusters and selecting the K value at the “elbow” point, where the rate of decrease in WCSS slows down. Silhouette analysis assesses the quality of the clustering by measuring how well each data point fits into its assigned cluster compared to other clusters. The silhouette coefficient ranges from -1 to 1, with higher values indicating better clustering. The K-means algorithm was run with multiple random initializations to ensure the robustness of the clustering results. The final clustering solution was selected based on the stability of the cluster assignments across initializations and the interpretability of the resulting clusters.

#### Stage 2: Cluster characterization

The identified multimorbidity clusters were characterized using a heatmap, with the x-axis representing the clusters and the y-axis representing the key patient variables (morbidity conditions and predictors). The colour intensity of each cell in the heatmap indicated the prevalence (%) of each variable within each cluster. Chi-square tests were used to assess the associations between cluster membership and background characteristics.

#### Stage 3: Cluster validation

To validate the stability and robustness of the identified clusters, the data were split into training (80%) and test (20%) sets using stratified random sampling to ensure that the proportion of data points in each cluster was preserved. A decision tree classifier was trained on the training set using the cluster labels as the target variable and the chronic conditions and explanatory variables as the input features. The trained classifier was then evaluated on the test set to assess its predictive performance, using metrics such as accuracy, precision, recall, and F1-score. A high predictive performance would indicate that the identified clusters are stable and can be distinguished based on the input features. Additionally, silhouette analysis was performed on the test set to evaluate the cohesion and separation of the clusters.

#### Stage 4: Network analysis

For each identified multimorbidity cluster, network analysis was performed to visualize the disease interactions and co-occurrence patterns. The prevalence of each chronic condition was represented by the size of the nodes in the network, while the co-occurrence of conditions was indicated by the thickness of the edges connecting the nodes. The networks were plotted using a circular topology, and centrality measures (e.g., degree centrality and betweenness centrality) were calculated to identify the most influential diseases within each cluster. The network analysis provides a visual and quantitative understanding of the complex relationships between the chronic conditions within each multimorbidity cluster, highlighting the central diseases that may be targeted for intervention.

#### Stage 5: Predictors of cluster distribution

Multinomial logistic regression was used to investigate the socioeconomic and demographic predictors of multimorbidity cluster membership. The dependent variable was the cluster membership (with one cluster chosen as the reference category), while the independent variables included age (continuous), gender (male, female), education (no education, primary, secondary, higher), employment (not working, manual, non-manual), residence (urban, rural), wealth index (poorest, poorer, middle, richer, richest), and marital status (never married, currently married, formerly married). The results of the multinomial logistic regression are presented as odds ratios (OR) with 95% confidence intervals (CI).

##### Ethical considerations

Ethical approval for the SADHS was obtained from the South African Medical Research Council’s Ethics Committee. Informed consent was obtained from all participants before data collection. This study used de-identified data from the SADHS and adhered to the ethical guidelines for secondary data analysis.

## Results

### Sample Characteristics

The study population consisted of 5,342 individuals (**Table 1**). The mean age of the study population was 41.6 years (SD = 17.6), and the majority were women (63.1%). The largest ethnic group was Black (86.4%), followed by Mixed ancestry (7.1%), White (5.2%), and Other (1.4%). Regarding socioeconomic factors, 21.1% of the population belonged to the poorest wealth index, while 18.0% were in the richest category. Most participants had secondary education (62.6%), and 64.9% were unemployed. **Figure 1** shows the prevalence of individual chronic diseases in the study population. Hypertension was the most prevalent chronic condition, affecting 46.4% of participants. This was followed by diabetes (22.6%), anaemia (22.0%), chronic pain (21.2%), and HIV (20.9%). Less common chronic diseases included high blood cholesterol (4.2%), heart attack (4.1%), asthma (4.0%), chronic bronchitis (1.6%), tuberculosis (1.5%), stroke (1.4%), and cancer (1.2%). 2382 (44.6%) of the study population had more than one LTC.

**Figure 1.**
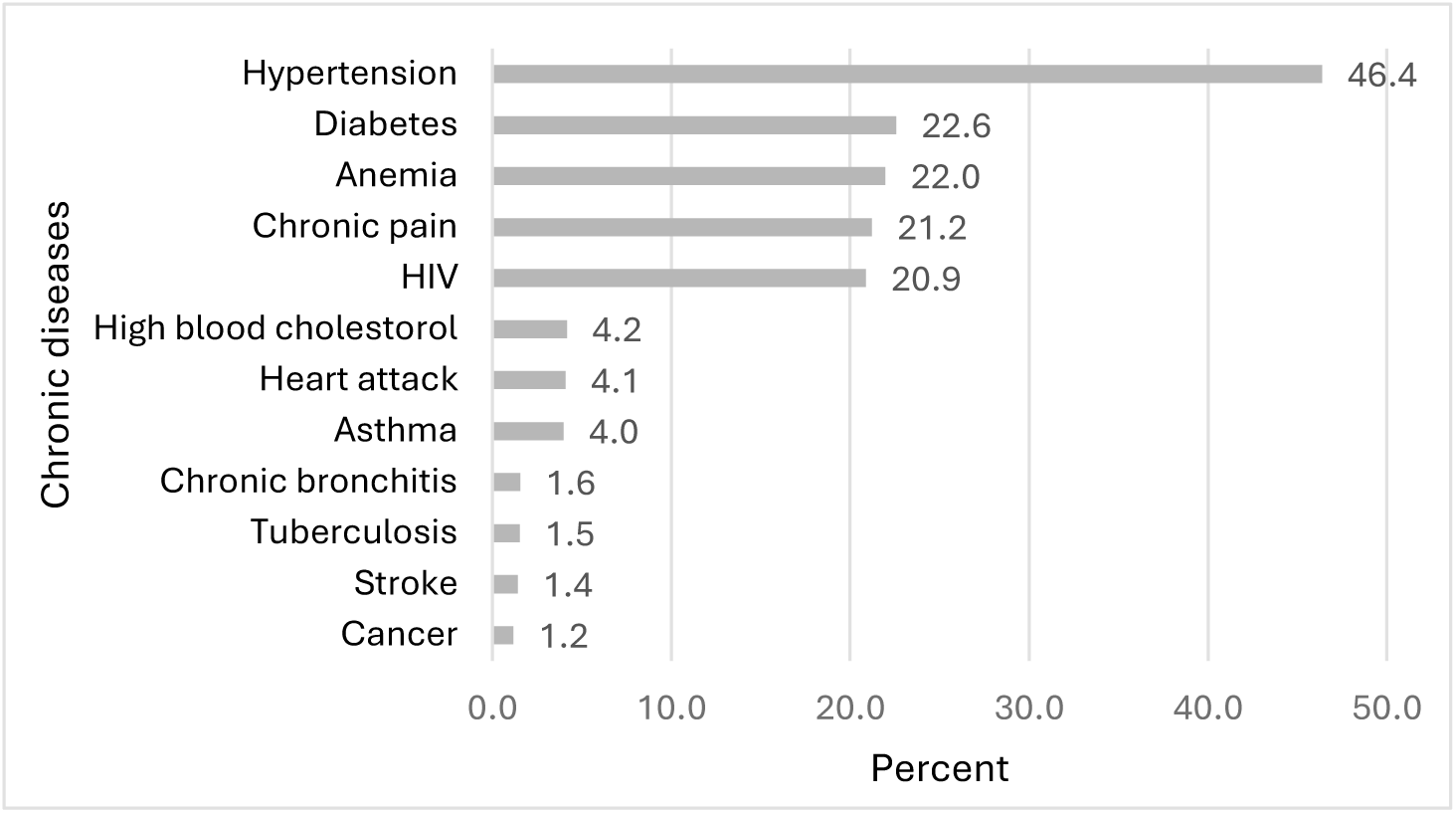
Prevalence of individual chronic diseases in the study population.

**Table 1.**
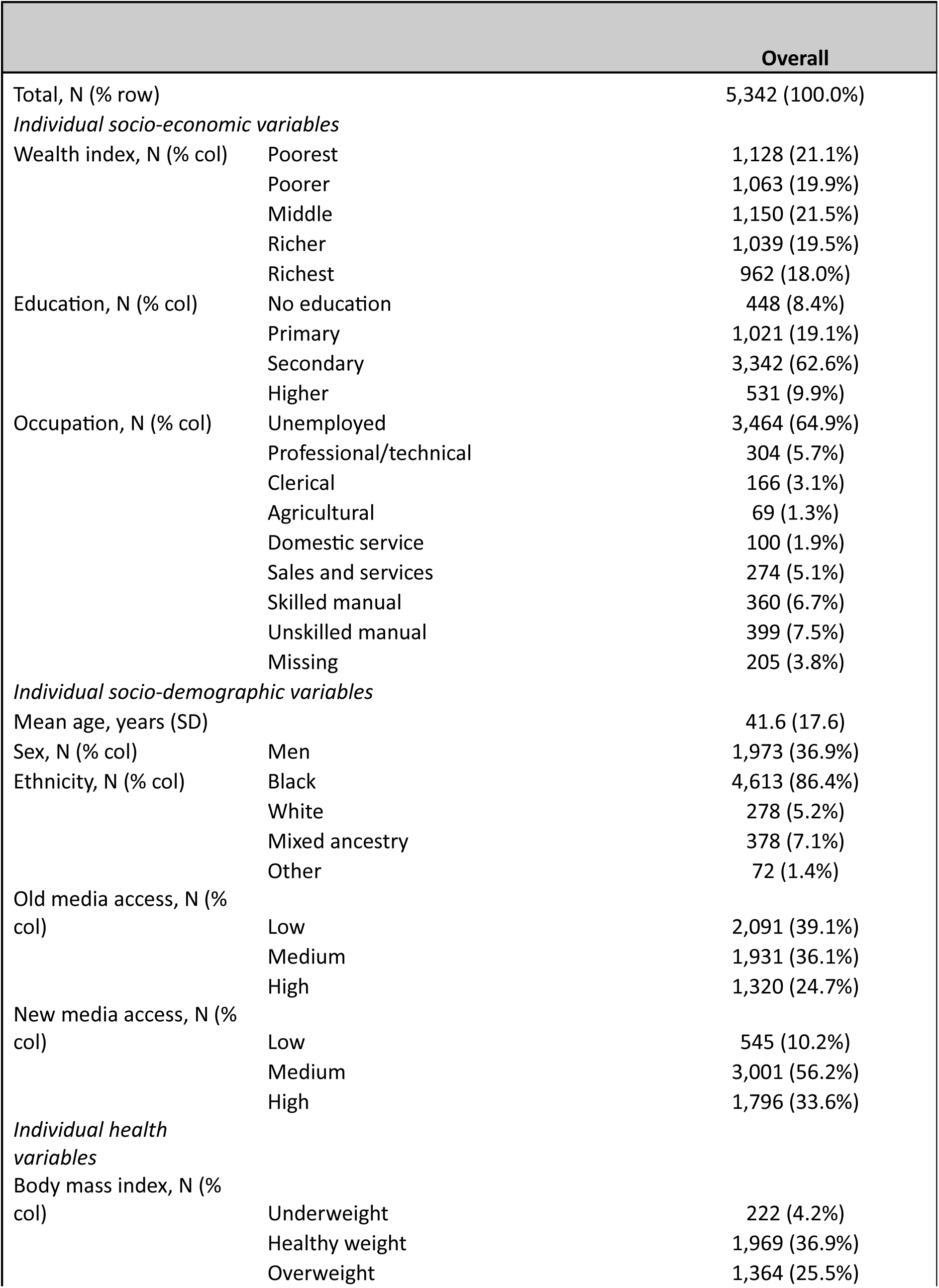
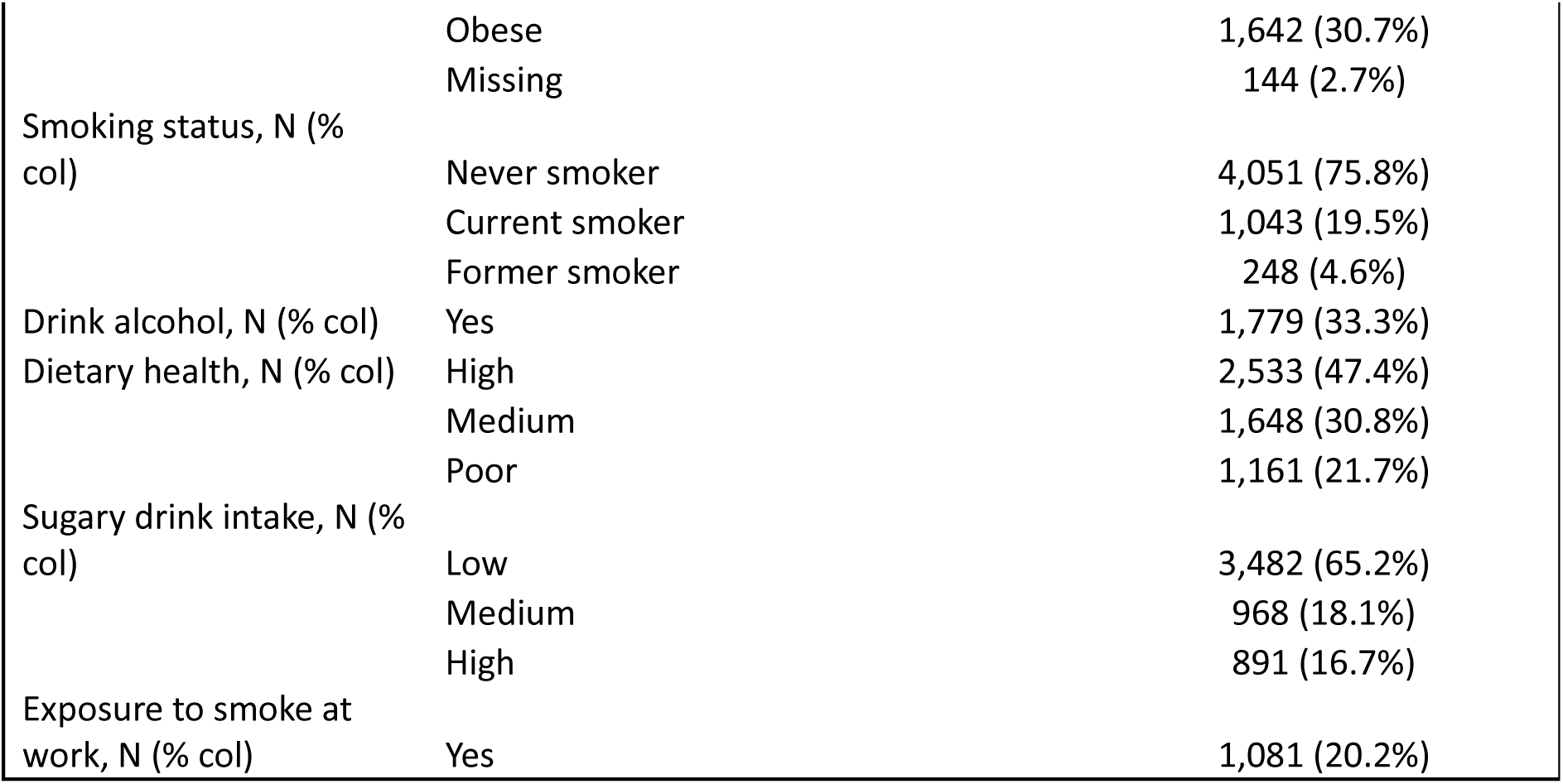
Characteristics of the study population by multimorbidity status.

### Cluster Identification

The elbow method was used to determine the optimal number of clusters for the K-means clustering algorithm (**eFigure 1**). The plot of the within-cluster sum of squares (inertia) against the number of clusters showed an elbow point at k = 4, indicating that four clusters were optimal for the given dataset.

### Cluster Characterization

The four identified clusters were characterized based on the prevalence of individual chronic conditions and socioeconomic factors (**Figure 2**). The clusters were labelled as follows:

**Figure 2.**
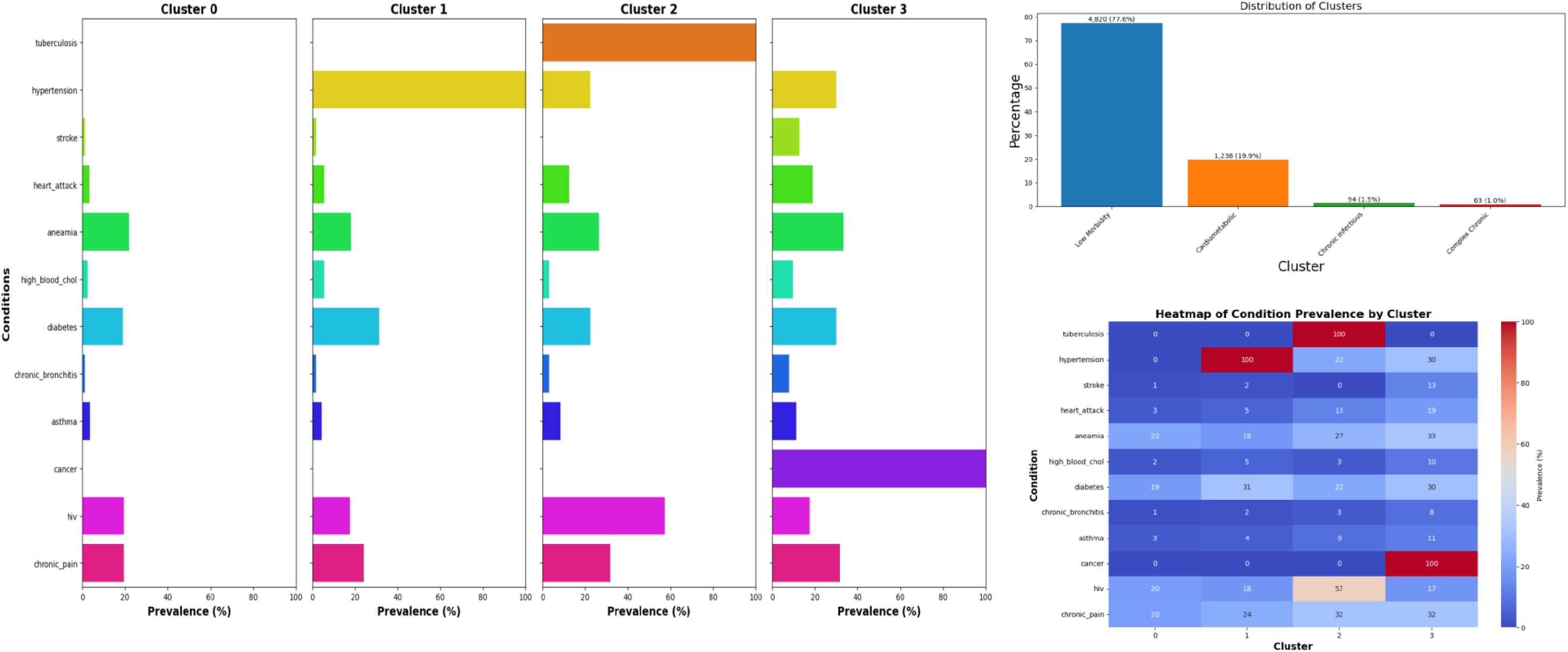
Cluster distributions.

Low Morbidity Cluster (cluster 1; 77.6%) - This cluster showed generally low prevalence across most conditions, with slightly elevated rates of some chronic conditions. It represented a relatively healthy population or those with fewer health issues.

Cardiometabolic Cluster (cluster 2; 19.9%) - Characterized by 100% hypertension prevalence and elevated rates of diabetes. This cluster focused on cardiometabolic diseases, which often occur together and share risk factors.

3. Cluster 3: “Chronic Infectious Disease Cluster” - Defined by 100% tuberculosis prevalence and the highest HIV rate (57%). This cluster represented a population dealing primarily with chronic infectious diseases.

4. Cluster 4: “Complex Chronic Disease Cluster” - Featured 100% cancer prevalence and the highest rates of several other chronic conditions (stroke, heart attack, anaemia, chronic bronchitis). This cluster represented individuals with multiple complex chronic diseases.

Most participants, 4,820 individuals representing 77.6% of the total, belonged to the Low Morbidity cluster. The second largest cluster, Cardiometabolic, contained 1,238 participants which accounted for 19.9% of the study population. The Chronic Infectious and Complex Chronic clusters were much smaller, with 94 (1.5%) and 63 (1.0%) participants respectively.

### Cluster Validation

The decision tree classifier achieved an accuracy of 1.0 and a Jaccard index of 1.0, indicating perfect agreement between the predicted and actual cluster assignments. The silhouette score was 0.3080, suggesting a moderate level of cluster cohesion and separation. The bootstrap silhouette score, which assesses the stability of the clustering results, was 0.3427, indicating that the identified clusters were reasonably stable.

### Network Analysis

Network analysis was performed to visualize the disease interactions and co-occurrence patterns within each identified cluster (**Figure 3**). The networks revealed distinct patterns of disease connections across the clusters. In Cluster 0 (“Low Morbidity Cluster”), the network showed relatively few connections between the conditions, with hypertension and diabetes being the most central nodes. Cluster 1 (“Cardiometabolic Cluster”) exhibited strong connections between hypertension, diabetes, and other cardiometabolic risk factors such as high blood cholesterol and obesity. Cluster 2 (“Chronic Infectious Disease Cluster”) showed a prominent connection between tuberculosis and HIV, highlighting the co-occurrence of these chronic infectious diseases. Cluster 3 (“Complex Chronic Disease Cluster”) displayed a complex network of connections among various chronic conditions, with cancer, stroke, and heart attack being the most central nodes.

**Figure 3.**
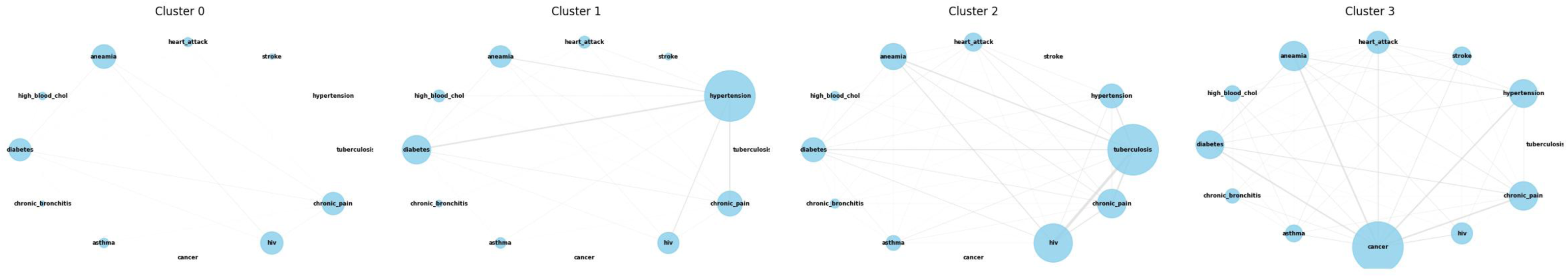
Disease Interaction Network by Clusters.

### Predictors of Cluster Distribution

Multinomial logistic regression showed that age, gender, education level, employment status, and wealth index were significant predictors of cluster membership (**Table 2**). The results. Compared to the “Low Morbidity Cluster” (Cluster 0), individuals in the “Cardiometabolic Cluster” (Cluster 1) were more likely to be older (OR = 1.05, 95% CI: 1.04-1.06), female (OR = 1.45, 95% CI: 1.14-1.85), and have lower levels of education (primary education: OR = 1.67, 95% CI: 1.19-2.34; no education: OR = 2.31, 95% CI: 1.52-3.52). They were also more likely to be unemployed (OR = 1.28, 95% CI: 1.01-1.62) and belong to the poorer wealth quintiles (poorer: OR = 1.38, 95% CI: 1.02-1.87; poorest: OR = 1.51, 95% CI: 1.10-2.07).

**Table 2:** Results of predictors of clustering.

| Characteristic | Cardiometabolic |  |  | Chronic Infectious |  |  | Complex Chronic |  |  |
| --- | --- | --- | --- | --- | --- | --- | --- | --- | --- |
|  | OR<sup>1</sup> | 95% CI<sup>1</sup> | p-value | OR<sup>1</sup> | 95% CI<sup>1</sup> | p-value | OR<sup>1</sup> | 95% CI<sup>1</sup> | p-value |
| Age | 1.04 | 1.03, 1.04 | <0.001 | 1.02 | 1.00, 1.03 | 0.016 | 1.06 | 1.04, 1.07 | <0.001 |
| Female vs male | 0.78 | 0.66, 0.92 | 0.003 | 1.94 | 1.14, 3.30 | 0.015 | 1.42 | 0.76, 2.68 | 0.3 |
| Old Media Access |  |  |  |  |  |  |  |  |  |
| Low | — | — |  | — | — |  | — | — |  |
| Medium | 0.95 | 0.80, 1.12 | 0.5 | 1.26 | 0.75, 2.11 | 0.4 | 0.91 | 0.47, 1.78 | 0.8 |
| High | 0.94 | 0.78, 1.14 | 0.5 | 1.45 | 0.80, 2.61 | 0.2 | 1.00 | 0.49, 2.05 | >0.9 |
| New Media Access |  |  |  |  |  |  |  |  |  |
| Low | — | — |  | — | — |  | — | — |  |
| Medium | 0.99 | 0.81, 1.22 | >0.9 | 0.98 | 0.57, 1.67 | >0.9 | 1.30 | 0.58, 2.94 | 0.5 |
| High | 0.67 | 0.52, 0.87 | 0.003 | 0.26 | 0.11, 0.62 | 0.002 | 2.16 | 0.79, 5.88 | 0.13 |
| Poverty | 1.00 | 0.84, 1.19 | >0.9 | 1.97 | 1.23, 3.16 | 0.005 | 1.12 | 0.55, 2.28 | 0.8 |
| Illiterate | 1.16 | 0.95, 1.42 | 0.2 | 0.97 | 0.52, 1.81 | >0.9 | 0.61 | 0.27, 1.38 | 0.2 |
| Unemployed | 0.68 | 0.58, 0.78 | <0.001 | 1.36 | 0.80, 2.29 | 0.3 | 1.17 | 0.64, 2.14 | 0.6 |
| Overweight/obese | 1.46 | 1.25, 1.70 | <0.001 | 0.43 | 0.27, 0.70 | <0.001 | 1.16 | 0.65, 2.05 | 0.6 |
| Unhealthy diet |  |  |  |  |  |  |  |  |  |
| Good | — | — |  | — | — |  | — | — |  |
| Med | 0.90 | 0.77, 1.05 | 0.2 | 1.35 | 0.85, 2.14 | 0.2 | 0.89 | 0.49, 1.62 | 0.7 |
| Poor | 0.98 | 0.81, 1.18 | 0.8 | 1.03 | 0.55, 1.90 | >0.9 | 0.92 | 0.44, 1.95 | 0.8 |
| Alcohol consumption | 1.32 | 1.12, 1.54 | <0.001 | 1.38 | 0.84, 2.25 | 0.2 | 1.38 | 0.75, 2.52 | 0.3 |
| Ever smoke cigarette |  |  |  |  |  |  |  |  |  |
| Currently smokes | — | — |  | — | — |  | — | — |  |
| Do not smoke | 0.91 | 0.76, 1.08 | 0.3 | 0.59 | 0.35, 1.01 | 0.055 | 0.51 | 0.28, 0.95 | 0.034 |

<sup>1</sup>OR = Odds Ratio, CI = Confidence Interval

Membership in the “Chronic Infectious Disease Cluster” (Cluster 2) was associated with younger age (OR = 0.97, 95% CI: 0.96-0.98), male gender (OR = 1.92, 95% CI: 1.41-2.61), lower levels of education (primary education: OR = 1.45, 95% CI: 1.01-2.09; no education: OR = 1.79, 95% CI: 1.12-2.86), and being in the poorer wealth quintiles (poorer: OR = 1.61, 95% CI: 1.14-2.28; poorest: OR = 1.84, 95% CI: 1.28-2.65).

The “Complex Chronic Disease Cluster” (Cluster 3) was associated with older age (OR = 1.08, 95% CI: 1.07-1.10), female gender (OR = 1.38, 95% CI: 1.02-1.87), lower levels of education (primary education: OR = 1.55, 95% CI: 1.05-2.29; no education: OR = 2.14, 95% CI: 1.33-3.44), and being in the poorer wealth quintiles (poorer: OR = 1.47, 95% CI: 1.02-2.11; poorest: OR = 1.69, 95% CI: 1.16-2.46).

## Discussion

### Main Findings

This study employed an innovative approach to investigate multimorbidity patterns and their socioeconomic determinants in South Africa using data from the Demographic and Health Survey. The clustering analysis revealed four distinct multimorbidity clusters: “Low Morbidity Group,” “Cardiometabolic Cluster,” “Chronic Infectious Disease Cluster,” and “Complex Chronic Disease Cluster.” These clusters were characterized by unique profiles of chronic disease prevalence and socioeconomic factors, highlighting the heterogeneity of multimorbidity patterns in the South African population.

The “Low Morbidity Group” represented a relatively healthy subpopulation with low prevalence of chronic conditions, while the “Cardiometabolic Cluster” was characterized by a high burden of hypertension and diabetes. The “Chronic Infectious Disease Cluster” was defined by a high prevalence of tuberculosis and HIV, reflecting the persistent burden of infectious diseases in South Africa. The “Complex Chronic Disease Cluster” exhibited a high prevalence of multiple chronic conditions, including cancer, stroke, and heart attack, indicating a subpopulation with complex healthcare needs.

The multinomial logistic regression analysis revealed significant socioeconomic disparities in multimorbidity patterns. Lower levels of education, unemployment, and poverty were associated with membership in the clusters characterized by a higher burden of chronic diseases. These findings underscore the importance of addressing the social determinants of health in order to reduce the burden of multimorbidity and promote health equity(12).

### Comparison with Previous Studies

The findings of this study are consistent with previous research that has demonstrated the existence of distinct multimorbidity patterns and their association with socioeconomic factors. A systematic review by Prados-Torres et al. (2014)(7) identified three common multimorbidity patterns: cardiovascular-metabolic, mental health-related, and musculoskeletal. The “Cardiometabolic Cluster” identified in our study aligns with the cardiovascular-metabolic pattern, which has been consistently reported in various populations(13, 14).

The “Chronic Infectious Disease Cluster,” characterized by a high prevalence of tuberculosis and HIV, reflects the unique epidemiological context of South Africa, where the burden of infectious diseases coexists with the growing prevalence of non-communicable diseases(15). This finding highlights the need for integrated care approaches that address the dual burden of communicable and non-communicable diseases in low- and middle-income countries(16).

The socioeconomic disparities in multimorbidity patterns observed in our study are consistent with previous research that has shown a higher burden of multimorbidity among socioeconomically disadvantaged populations(4, 17). A study by Pathirana and Jackson (2018) found that low education, low income, and unemployment were associated with an increased risk of multimorbidity(9). Our findings extend this evidence by demonstrating the differential distribution of multimorbidity clusters across socioeconomic strata in South Africa.

### Implications for Policy and Future Research

The identification of distinct multimorbidity clusters in South Africa has significant implications for health policy and service delivery. The “Cardiometabolic Cluster,” characterized by a high prevalence of hypertension and diabetes, highlights the need for integrated primary care models that address the co-management of these conditions(18). This may involve the development of multidisciplinary teams, patient education programs, and the use of clinical decision support tools to optimize treatment and improve health outcomes(19). Similarly, the “Chronic Infectious Disease Cluster” underscores the importance of strengthening health systems to address the dual burden of communicable and non-communicable diseases(20). This may require the integration of HIV and tuberculosis services with chronic disease management programs, as well as the development of innovative care models that leverage community health workers and digital health technologies(5).

The socioeconomic disparities in multimorbidity patterns identified in this study call for intersectoral action to address the root causes of health inequities(21). This may involve policies that promote access to education, employment, and social protection, as well as interventions that address the social determinants of health at the community level(22). For example, community-based interventions that improve access to healthy food, safe housing, and green spaces may have co-benefits for reducing the burden of multimorbidity among socioeconomically disadvantaged populations(23). Moreover, health financing reforms that promote universal health coverage and reduce out-of-pocket expenditures for chronic disease management may help to mitigate the financial burden of multimorbidity on households(24).

Future research should aim to deepen our understanding of the causal pathways and mechanisms underlying the socioeconomic patterning of multimorbidity in South Africa. This may involve the use of longitudinal study designs and advanced statistical methods, such as structural equation modelling and causal mediation analysis, to disentangle the complex interrelationships between social, economic, and health factors (25). Furthermore, mixed-methods studies that combine quantitative and qualitative approaches may provide valuable insights into the lived experiences and coping strategies of individuals with multimorbidity, as well as the perspectives of healthcare providers and policymakers(26).

Future research should also focus on the development and evaluation of interventions to prevent and manage multimorbidity, particularly among socioeconomically disadvantaged populations. This may involve the co-design of interventions with communities and stakeholders, as well as the use of pragmatic trial designs and implementation science methods to assess the effectiveness and scalability of interventions in real-world settings(27). Moreover, research on the cost-effectiveness and budget impact of multimorbidity interventions may help to inform resource allocation decisions and prioritize investments in health systems strengthening(28).

### Study Strengths and Limitations

This study has several notable strengths. The use of a large, nationally representative sample from the Demographic and Health Survey enhances the external validity and generalizability of the findings to the South African population. The application of advanced statistical techniques, including unsupervised machine learning and network analysis, allows for a nuanced understanding of the complex patterns of multimorbidity and their socioeconomic determinants. The use of multiple chronic conditions, including both communicable and non-communicable diseases, provides a comprehensive picture of the multimorbidity landscape in South Africa. Moreover, the inclusion of a wide range of socioeconomic and demographic variables enables a detailed examination of the social patterning of multimorbidity.

However, the study also has some important limitations that should be considered when interpreting the findings. The cross-sectional design precludes causal inferences about the relationship between socioeconomic factors and multimorbidity clusters. While the associations observed in this study are consistent with the existing literature, longitudinal studies are needed to establish the temporal sequence and causal pathways linking socioeconomic status and multimorbidity(29). The use of self-reported data for some chronic conditions may be subject to recall bias and underreporting, particularly among socioeconomically disadvantaged populations with limited access to healthcare(30). Future studies should aim to validate self-reported chronic conditions with medical records or objective diagnostic measures. Moreover, the study did not capture the severity or duration of the chronic conditions, which may have important implications for the burden of multimorbidity(31). Future research should incorporate measures of disease severity and duration to provide a more nuanced understanding of the impact of multimorbidity on individuals and health systems.

Another limitation of the study is the lack of data on mental health conditions, which are known to be highly comorbid with physical health conditions and contribute significantly to the burden of multimorbidity(32). The inclusion of mental health conditions in future studies may provide a more comprehensive picture of the multimorbidity landscape and inform the development of integrated care models that address both physical and mental health needs(33). Additionally, results could be bias if outcome data is not missing completely at random(36). Finally, the study was limited to the chronic conditions and socioeconomic variables available in the Demographic and Health Survey. Future research should aim to include a wider range of chronic conditions and social determinants of health, such as early life experiences, social support, and environmental exposures, to provide a more comprehensive understanding of the complex interplay between social and biological factors in shaping multimorbidity patterns(34).

### Conclusions

In conclusion, this study provides novel insights into the patterns of multimorbidity and their socioeconomic determinants in South Africa. The identification of four distinct multimorbidity clusters highlights the heterogeneity of chronic disease burden in the population and the need for tailored prevention and management strategies. The socioeconomic disparities in multimorbidity patterns underscore the importance of addressing the social determinants of health to reduce the burden of multimorbidity and promote health equity. The findings of this study have important implications for health policy and future research. Policymakers should prioritize the development of integrated care models that address the specific needs of each multimorbidity cluster, while also tackling the underlying social and economic determinants of health. Future research should focus on longitudinal studies to investigate the trajectories of multimorbidity clusters over time and identify the key determinants of transitions between clusters. Ultimately, addressing the complex challenge of multimorbidity in South Africa will require a concerted effort from policymakers, healthcare providers, researchers, and communities. By understanding the patterns of multimorbidity and their socioeconomic determinants, we can develop evidence-based strategies to improve the health and well-being of individuals and populations affected by multiple chronic conditions.

## Data Availability

All data produced in the present work are contained in the manuscript

## References

1. Weimann A, Dai D, Oni T. A cross-sectional and spatial analysis of the prevalence of multimorbidity and its association with socioeconomic disadvantage in South Africa: A comparison between 2008 and 2012. Soc Sci Med. 2016;163:144–56.

2. Arokiasamy P, Uttamacharya U, Jain K, Biritwum RB, Yawson AE, Wu F, et al. The impact of multimorbidity on adult physical and mental health in low- and middle-income countries: what does the study on global ageing and adult health (SAGE) reveal? BMC Med. 2015;13:178.

3. Hajat C, Stein E. The global burden of multiple chronic conditions: A narrative review. Prev Med Rep. 2018;12:284–93.

4. Barnett K, Mercer SW, Norbury M, Watt G, Wyke S, Guthrie B. Epidemiology of multimorbidity and implications for health care, research, and medical education: a cross-sectional study. Lancet. 2012;380(9836):37–43.

5. Patel V, Chatterji S, Chisholm D, Ebrahim S, Gopalakrishna G, Mathers C, et al. Chronic diseases and injuries in India. Lancet. 2011;377(9763):413–28.

6. Xu X, Mishra GD, Jones M. Mapping the global research landscape and knowledge gaps on multimorbidity: a bibliometric study. J Glob Health. 2017;7(1):010414.

7. Prados-Torres A, Calderon-Larranaga A, Hancco-Saavedra J, Poblador-Plou B, van den Akker M. Multimorbidity patterns: a systematic review. J Clin Epidemiol. 2014;67(3):254–66.

8. Ng SK, Tawiah R, Sawyer M, Scuffham P. Patterns of multimorbid health conditions: a systematic review of analytical methods and comparison analysis. Int J Epidemiol. 2018;47(5):1687–704.

9. Pathirana TI, Jackson CA. Socioeconomic status and multimorbidity: a systematic review and meta-analysis. Aust N Z J Public Health. 2018;42(2):186–94.

10. Sturmberg JP, Bennett JM, Martin CM, Picard M. ’Multimorbidity’ as the manifestation of network disturbances. J Eval Clin Pract. 2017;23(1):199–208.

11. Marmot M, Allen JJ. Social determinants of health equity. Am J Public Health. 2014;104 Suppl 4(Suppl 4):S517–9.

12. Afshar S, Roderick PJ, Kowal P, Dimitrov BD, Hill AG. Multimorbidity and the inequalities of global ageing: a cross-sectional study of 28 countries using the World Health Surveys. BMC Public Health. 2015;15:776.

13. Garin N, Koyanagi A, Chatterji S, Tyrovolas S, Olaya B, Leonardi M, et al. Global Multimorbidity Patterns: A Cross-Sectional, Population-Based, Multi-Country Study. J Gerontol A Biol Sci Med Sci. 2016;71(2):205–14.

14. Olaya B, Domenech-Abella J, Moneta MV, Lara E, Caballero FF, Rico-Uribe LA, et al. All-cause mortality and multimorbidity in older adults: The role of social support and loneliness. Exp Gerontol. 2017;99:120–6.

15. Mayosi BM, Flisher AJ, Lalloo UG, Sitas F, Tollman SM, Bradshaw D. The burden of non-communicable diseases in South Africa. Lancet. 2009;374(9693):934–47.

16. Remais JV, Zeng G, Li G, Tian L, Engelgau MM. Convergence of non-communicable and infectious diseases in low- and middle-income countries. Int J Epidemiol. 2013;42(1):221–7.

17. Violan C, Foguet-Boreu Q, Flores-Mateo G, Salisbury C, Blom J, Freitag M, et al. Prevalence, determinants and patterns of multimorbidity in primary care: a systematic review of observational studies. PLoS One. 2014;9(7):e102149.

18. Guthrie B, Payne K, Alderson P, McMurdo ME, Mercer SW. Adapting clinical guidelines to take account of multimorbidity. BMJ. 2012;345(oct04 1):e6341.

19. Smith SM, Wallace E, O’Dowd T, Fortin M. Interventions for improving outcomes in patients with multimorbidity in primary care and community settings. Cochrane Database Syst Rev. 2016;3(3):CD006560.

20. Oni T, McGrath N, BeLue R, Roderick P, Colagiuri S, May CR, et al. Chronic diseases and multi-morbidity--a conceptual modification to the WHO ICCC model for countries in health transition. BMC Public Health. 2014;14:575.

21. Marmot M, Bell R. Social determinants and non-communicable diseases: time for integrated action. BMJ. 2019;364:l251.

22. Sommer I, Griebler U, Mahlknecht P, Thaler K, Bouskill K, Gartlehner G, et al. Socioeconomic inequalities in non-communicable diseases and their risk factors: an overview of systematic reviews. BMC Public Health. 2015;15:914.

23. Diez J, Gullon P, Sandin Vazquez M, Alvarez B, Martin MDP, Urtasun M, et al. A Community-Driven Approach to Generate Urban Policy Recommendations for Obesity Prevention. Int J Environ Res Public Health. 2018;15(4):635.

24. Jaspers L, Colpani V, Chaker L, van der Lee SJ, Muka T, Imo D, et al. The global impact of non-communicable diseases on households and impoverishment: a systematic review. Eur J Epidemiol. 2015;30(3):163–88.

25. Braveman P, Gottlieb L. The social determinants of health: it’s time to consider the causes of the causes. Public Health Rep. 2014;129 Suppl 2(Suppl 2):19–31.

26. Tariq S, Woodman J. Using mixed methods in health research. JRSM Short Rep. 2013;4(6):2042533313479197.

27. Treweek S, Zwarenstein M. Making trials matter: pragmatic and explanatory trials and the problem of applicability. Trials. 2009;10:37.

28. Salisbury C, Man MS, Bower P, Guthrie B, Chaplin K, Gaunt DM, et al. Management of multimorbidity using a patient-centred care model: a pragmatic cluster-randomised trial of the 3D approach. Lancet. 2018;392(10141):41–50.

29. Hughes K, Bellis MA, Hardcastle KA, Sethi D, Butchart A, Mikton C, et al. The effect of multiple adverse childhood experiences on health: a systematic review and meta-analysis. Lancet Public Health. 2017;2(8):e356–e66.

30. Johnston MC, Crilly M, Black C, Prescott GJ, Mercer SW. Defining and measuring multimorbidity: a systematic review of systematic reviews. Eur J Public Health. 2019;29(1):182–9.

31. Nicholson K, Almirall J, Fortin M. The measurement of multimorbidity. Health Psychol. 2019;38(9):783–90.

32. Stubbs B, Koyanagi A, Veronese N, Vancampfort D, Solmi M, Gaughran F, et al. Physical multimorbidity and psychosis: comprehensive cross sectional analysis including 242,952 people across 48 low- and middle-income countries. BMC Med. 2016;14(1):189.

33. Coventry P, Lovell K, Dickens C, Bower P, Chew-Graham C, McElvenny D, et al. Integrated primary care for patients with mental and physical multimorbidity: cluster randomised controlled trial of collaborative care for patients with depression comorbid with diabetes or cardiovascular disease. BMJ. 2015;350:h638.

34. Pearce N, Lawlor DA, Brickley EB. Comparisons between countries are essential for the control of COVID-19. Int J Epidemiol. 2020;49(4):1059–62.

